# Mathematical model study of a pandemic: Graded lockdown approach

**DOI:** 10.1101/2020.07.22.20159962

**Authors:** S. Chatterjee, V.C. Vani, Ravinder K. Banyal

## Abstract

A kinetic approach is developed, in a “tutorial style” to describe the evolution of an epidemic with spread taking place through contact. The “infection - rate” is calculated from the rate at which an infected person approaches an uninfected susceptible individual, i.e. a potential recipient of the disease, up to a distance *p*, where the value of *p* may lie between *p*_*min*_ *≤ p ≤ p*_*max*_. We consider a situation with a total population of *N* individuals, living in an area *A, x(t)* amongst them being infected while *x*_*d*_*(t) = β′x(t)* is the number that have died in the course of transmission and evolution of the epidemic. The evolution is developed under the conditions (1) a faction α*(t)* of the *[N-x(t) – x*_*d*_*(t)]* uninfected individuals and (2) a *β(t)* fraction of the *x(t)* infected population are quarantined, while the “source events” that spread the infection are considered to occur with frequency *υ*_*0*_. The processes of contact and transmission are considered to be Markovian. Transmission is assumed to be inhibited by several processes like the use of “masks”, “hand washing or use of sanitizers” while “physical distancing” is described by *p*. The evolution equation for *x*(*t*) is a Riccati - type differential equation whose coefficients are time-dependent quantities, being determined by an interplay between the above parameters. A formal solution for *x(t)* is presented, for a *“graded lockdown”* with the parameters, *0≤ α(t), β(t)≤1* reaching their respective saturation values in time scales, *τ*_*1*_, *τ*_*2*_ respectively, from their initial values *α(0)=β(0)=0*. The growth is predicted for several BBMP wards in Bengaluru and in urban centers in Chikkaballapur district, as an illustrative case. Above selections serve as model cases for high, moderate and thin population densities. It is seen that the evolution of *[x(t)/N]* with time depends upon (a) the initial time scale of evolution, (b) the time scale of cure and (c) on the time dependence of the Lockdown function *Q(t) = {[1-α(t)]·[1-β(t)]}*. The formulae are amenable to simple computations and show that in order to curb the spread one must ensure that *Q(∞)* must be below a critical value and the vigilance has to be continued for a long time (at least 100 to 150 days) after the decay starts, to avoid all chances of the infection reappearing.

## Introduction

Unprecedented global spread of Covid-19 pandemic has exposed our vulnerability. Ever since the Chinese declared, on 8^th^ January 2020, transmission of infection due to this highly communicable virus, various steps have been contemplated and executed to “break-the-chain” of Covid-19’s transmission, particularly after the WHO’s declaration of this virus to be a cause of public emergency (31^st^ January, 2020) and a pandemic (11^th^ March, 2020). Thanks to very rapid information flow on-line, people are aware of different control strategies, which can be: (A). Aggressive complete lockdown, meant to beat the virus’s growth and flattening the curve in mere few weeks; or, (B) Selective and smart containment by proper detection of infected persons and quarantining them urgently and (C) Contact tracing of the primary and secondary contacts of the infected and isolating them to avoid further spread.

While we are flooded with figures, graphs and charts these are overall national level figures. The epidemic however, does not spread “nationally” in a uniform way. It first spreads in local areas, spills over and expands the boundaries of its “operation”. Containment strategies are to be local, as is now recognized. Thanks to awareness campaigns by several agencies, universal use of masks by all citizens, quarantine of infected people and the risk of infection from asymptomatic cases are now part of public knowledge. However, people and most likely local level planners are still unaware of a way to predict the progress of the spread, when an infection gets planted in their own area. Also, people’s concerns are, “how long do we have to wait to get rid of the red or danger zone label and be declared an orange one and then as a green or safe zone? How long should we keep our activities on hold? Who should move and who should be quarantined?” A local level planner must be able to calculate the upper bounds, in order to be ready with a realistic scenario and plan the strategy.

The present paper addresses these questions, through a <u>logistic rate process</u> approach [1-3], developed from the first principles, on the basis of kinetics of encounter. Firstly, we assume that the confinement strategy is implemented municipal ward wise and movements are restricted within a ward so that leakage of infection does neither flow out, nor it is imported from outside. It thus does NOT address the result of nationwide lockdown but limits itself to a more elementary but fundamental problem of ward-wise confinement. The rate at which the cases can grow within the ward are determined by, (a) population density *(ρ)*, (b) infective time-scale *(τ*_*0*_), (c) the quarantine coefficients *(α(t), β(t))* which are the fractions of uninfected and infected people that are quarantined, (d) the recovery times *(τ*_*1*_, *τ*_*2*_) of the quarantined and un-quarantined infected people.

Demand of “exactness” puts insurmountable obstacles before theorists, with availability of reliable data being a barrier. Also, trying for grand “national” scenario is another, which demands that all the possibilities have to be incorporated to make models ready-made for “one fits all”. Neither of these is attempted here. The objective is to understand the evolution of a disease in a limited sphere, based on an understanding that the spread would depend upon (a) encounter between infected and uninfected individuals within the “infective distance*”(p*_*0*_), (b) strength of the infective process, i.e. if people are wearing masks. These processes would determine the initial time scale of growth *(τ*_*0*_). How the evolution of the infection would unfold would be determined by (c) how fast the quarantining factors *(α(t), β(t))* grow and (d) how quickly does the epidemic’s growth get arrested due to recovery time-scales *(τ*_*1*_, *τ*_*2*_).

The numbers used here are illustrative of typical processes, of different municipal wards of Bruhat Bengaluru Mahanagara Palike (BBMP). Population data are obtained from BBMP website and report of spread from newspapers. Typical population in a BBMP ward is 30,000-40,000 and the area can be 0.31-1.0 square-kilometers in most cases and hence the population density can range from 10,00-120,00 per sq.km. In smaller towns outside the metropolis, the typical density of population can be 5000-15000 per square kilometer. Growth rate predictions are made for some typical cases in BBMP wards and towns in Chikkaballapur district.

## Theory

In the proposed SIR (susceptible-infected-replacement) [1-3] model with graded lockdown, let us consider:

*N*= total number of individuals in a population

*A*= area of their residence

*ρ= [N/A] =* number density of population

*x(t) =* number of infected persons

*x*_*d*_*(t)* = *β′x(t)* = number died, where a proportionality is assumed

*α(t)* = fraction of uninfected people who are quarantined

*1-α(t)* = fraction of uninfected people who are not quarantined

*β(t)* = fraction of infected people who are quarantined

*1-β(t)* = fraction of infected people who are not quarantined

*N′(t) = N-x(t)-x*_*d*_*(t) = N-x(t) – β′ x(t) = N – (1+β′)x(t) =* total number of people who are not infected

*ρ′ = N’/A = ρ [1-x(t)/N – x*_*d*_*(t)/N] = ρ [1- (1 + β′)x(t)] =* density of uninfected people.

***v ≡*** *(v*_*x*_, *v*_*y*_*) =* velocity of infected individuals and the components of the velocities.

***v′ ≡*** *(v′*_*x*_,*v′*_*y*_*) =* velocity of non-infected individuals and the components of the velocities

*f(v*_*x*_, *v*_*y*_*) dv*_*x*_ *dv*_*y*_ = probability of finding infected individuals in the velocity range *(v*_*x*_, *v*_*y*_) and *(v*_*x*_ *+ dv*_*x*_, *v*_*y*_ *+ dv*_*y*_ )

*f(v′*_*x*_, *v′*_*y*_*) dv′*_*x*_ *dv′*_*y*_*) =* probability of finding infected individuals in the velocity range *(v′*_*x*_, *v′*_*y*_) and *(v′*_*x*_ *+ dv′*_*x*_, *v′*_*y*_ *+ dv′*_*y*_ )

*τ*_*1*_ = recovery time for the quarantined cases

*τ*_*2*_ = recovery time for the non-quarantined cases.

In terms of the existing nomenclature in the literature, we have the number of infected people to be *I= x(t)* and the susceptible number is, *S=N′(t)*, which is directly expressed in terms of *I*. For describing the growth, [1-3] we follow a kinetic approach, i.e. find a rate of encounter between the *I* and *S* sections of the population [4-5]. Both the processes are stochastic, which finally lead us to a logistic equation of growth by keeping in mind that the capacity of an encounter to pass on infection is curbed by (a) quarantining a fraction *α(t)* of the *S* section of the population and (b) a fraction *β(t)* of the *I* section of the population and by (c) “*social vaccination*”, like the use of masks. The quarantine factors, *α(t)* and *β(t)* are time dependent, describing a graded lockdown for containment strategy. The present model considers evolution in absence immunization by either herd immunity or by vaccination. The population *R* is removed from the susceptible section by death, i.e. *R = x*_*d*_ *(t) = β′x(t)*. This evolution process would thus yield an overestimate, but nevertheless can act as a guide by giving *upper bounds* for the rate of growth and saturation.

The model considers the growth process in a restricted region of area *A* and considers that un-quarantined are free to move in this region. Also, under this containment strategy, the un-quarantined *I* component of the population infect only people within this zone. Once the containment starts there is no entry of infection from outside, nor migrations outside.

### Dynamics of “approach” between the infected and uninfected

We assume that the infected and the uninfected move at random in memory-less Markoffian fashion, so that any encounter is independent of the history of past interactions. Further, in the present case, approach towards one individual does not alter the path of the encounter. Referring to Figure 1, let us consider any particular trajectory and let *p* be the distance of closest approach between the infected and uninfected individuals in a trajectory, to be called the *“impact parameter”* of the trajectory.

**Figure 1.**
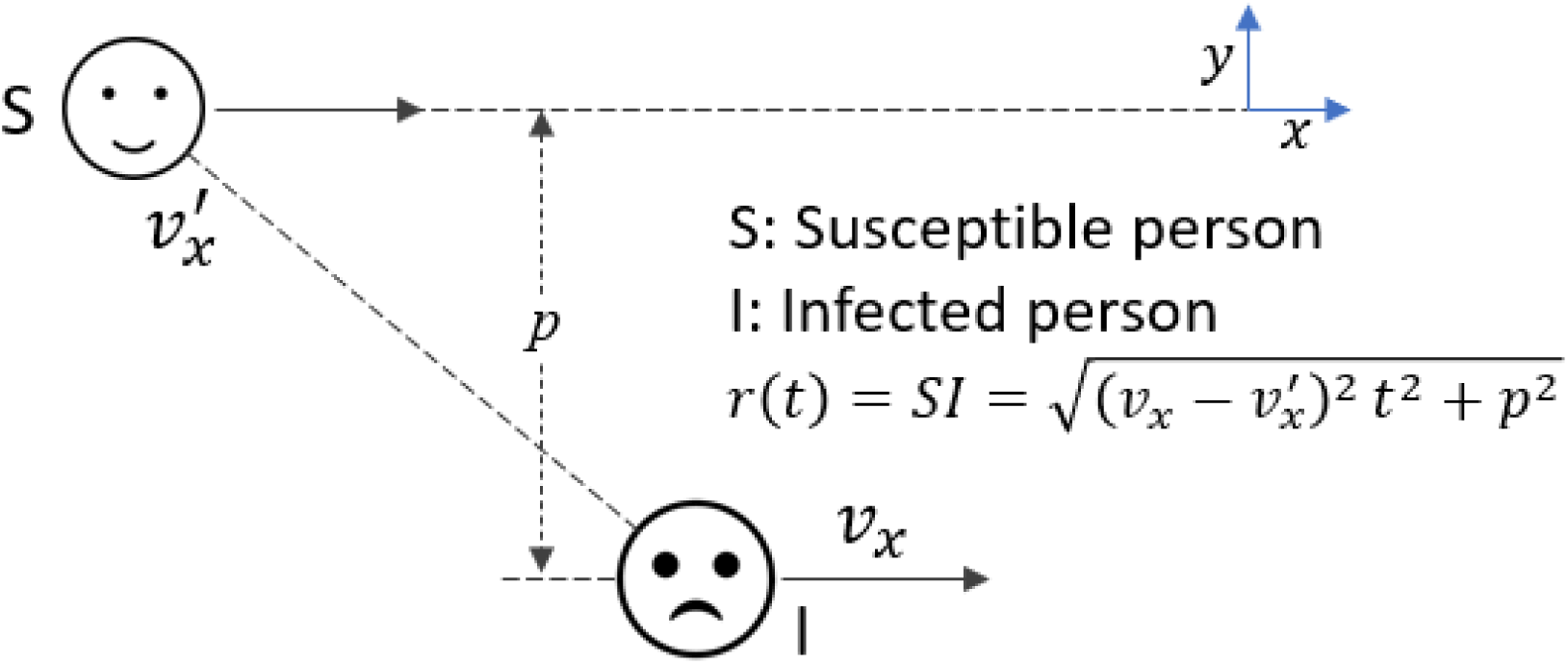
Geometry of encounter between the infected person (I) and the susceptible person (S), with their velocities being *v*_*x*_ and *v*_*x*_*′*, respectively in the x-direction, with *p* being the impact parameter.

Let us consider an infected non-quarantined individual (I, in the figure 1), who is traveling with a velocity *v*_*x*_ in the x-direction and let us consider his/her encounters with uninfected people (S, in the figure 1), located within impact parameter range *p* and *(p+δp*) in a time interval *δt*, where uninfected individuals are traveling with velocities in the range velocity range *(v′*_*x*_, *v′*_*y*_) and *(v′*_*x*_ *+ dv′*_*x*_, *v′*_*y*_ *+ dv′*_*y*_ ). Thus, all the non-quarantined non-infected individuals located within the area | *v*_*x*_ *-v′*_*x*_ | *δt δp*, will experience “encounters” with this infected individual in time *δt* and have a chance of infection. This number is number of encounters in time *δt* is:

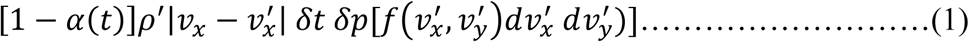

Since the number of such non quarantined infected individuals is *[1-β(t)]x(t)*, the number of such potential infective encounters in time *δt* is given by:

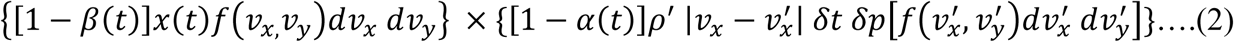

To determine the increase in the number of infected cases, we have to multiply (2) by a factor: *η*, which is the average number of infective events that take place per collision.

## Calculation of η

Any encounter extends from *t =* − *∞* to *t = + ∞*, during which the infective individual lies at *x(t) =* |*v*_*x*_ *−v′*_*x*_ |(*t,p*), i.e. at a distance,

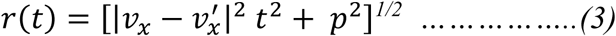

from the susceptible one. We consider that the strength of the infection falls off monotonically as *S(r(t))*, with the distance *r(t)* from the source of infection. Let [υ_0_ *δt*] be the number of infective events (e.g. cough, sneeze etc.) that take place in time interval *δt*, so that in an encounter that extends between *t = − ∞* to *t = + ∞*, the effective strength of infection is

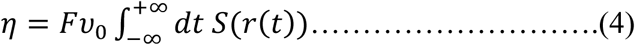

where *F ≤* 1, is the *“protection factor”* arising out of use of masks etc.

## Models for S(r(t))

### 1. Exponential case

Let

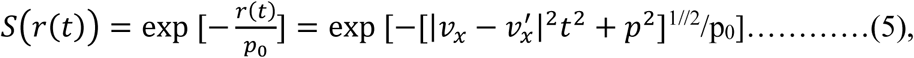

where *p*_*0*_ is a scale length beyond which the strength of imparting infection drops rapidly. Then, on integration, we find,

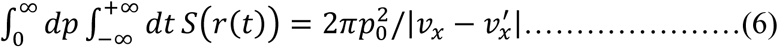

### 2. Gaussian case

Let

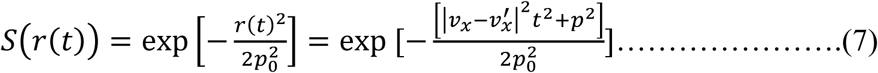

then,

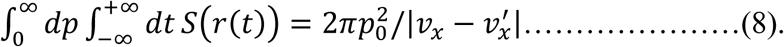

### 3. Lorentzian case

Let,

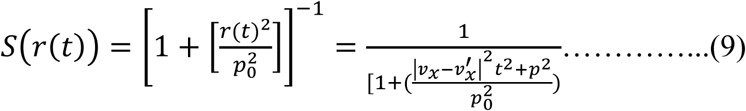

then,

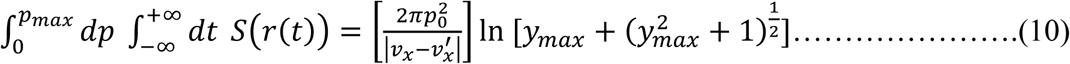

With

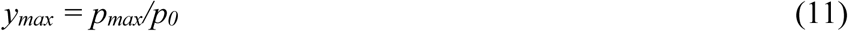

where, we have to put an upper value *p*_*max*_ for the impact parameter, since otherwise the integral over *p* diverges. This is because the Lorentzian function falls too slowly, giving rise to a logarithmic divergence. Such divergences do not take place in the exponential and Gaussian cases, since they fall rapidly.

In general, we can write,

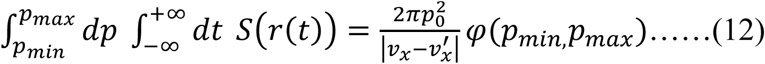

The expressions in (6), (8), (11) would be smaller those given, since the lower limit of integration over *p* is, *p*_*min*_ *>0*, though we have taken it to be zero, both for simplifying the results and also because physical distancing is not generally enforced and is also not practical for several circumstances.

Then the expression in (2) reads, after performing the integration over *p*,

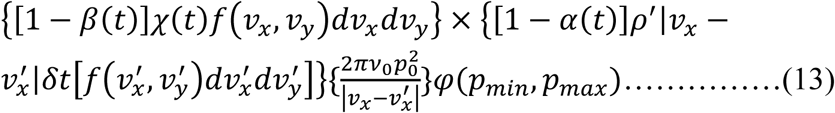

We notice that | *v*_*x*_ *−v′*_*x*_ | being present in both numerator and denominator of (13), they cancel each other. This may appear surprising, since infection transmission would thus arise, even if individuals do not encounter each other! The idea can be resolved as follows. The flux of “collision” is proportional to *∼*| *v*_*x*_ *−v′*_*x*_ |, but the time to move away, beyond the “infection scale length” *p*_*0*_, goes as *t*_*coll*_ *∼ p*_*0*_*/*|*v*_*x*_ *-v′*_*x*_ |, within which time *[υ*_*0*_ *t*_*coll*_ ] infective events take place. Thus, the net rate of imparting infection would go as, *[flux]×[υ*_*0*_ *t*_*coll*_ *] ∼ [υ*_*0*_*p*_*0*_ ], which is independent of | *v*_*x*_ *-v*′_*x*_ |.

Thus, on noting that

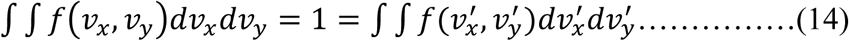

we can from (17) on making *δt* →*0*, get the rate of increase of infected cases, due to encounter. To take care of other factors that inhibit infection, we include the following multiplicative factors.

1. Geometrical Exposure Factor

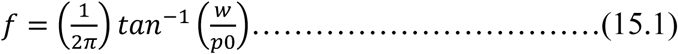

since the infection event can succeed only if the “victim” stands face to face with the infected person, i.e. within and angle tan^-1^(w/p_0_).
2. Protective factors *f*_*1*_ *≤ 1* and *f*_*2*_ *≤* 1, which describe diminutions of the power of infection due to usage of masks by the infected person and uninfected person. The net effect would be a factor

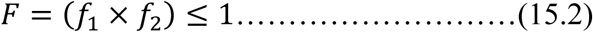

Thus, on multiplying (17) by *[f*.*F]* and letting *δt* → *0*, we get the logistic equation of growth due to encounter to be,

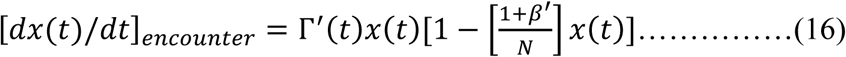

With

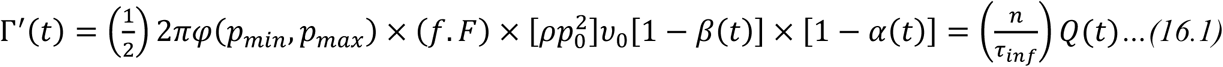

where,

*n = [2π ρp*_*0*_^2^] = average number of people in the infective circle

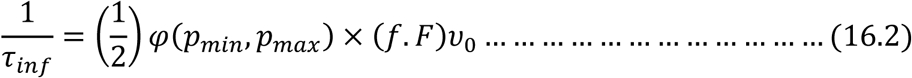

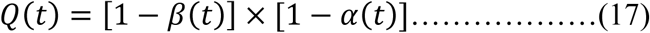

where, in defining τ_inf_ as a typical time scale for transmission of infection, a factor of *(1/2)* is incorporated since people do not move around at night, i.e. for half the day.

The decrement in the infected cases, due to recovery goes as,

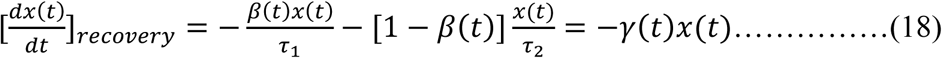

and the decrement due to death is given by,

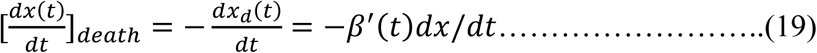

Thus, the total rate of change is given by,

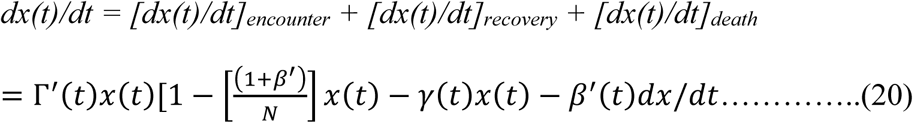

i.e.

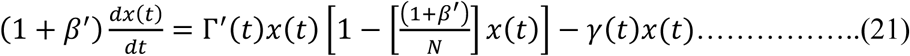

This finally gives the evolution to follow,

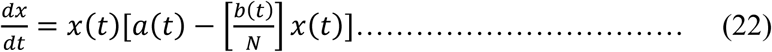

where,

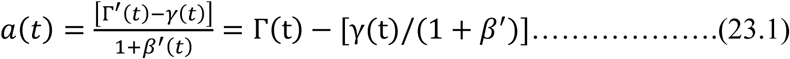

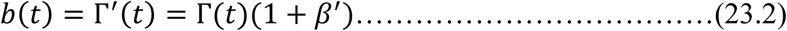

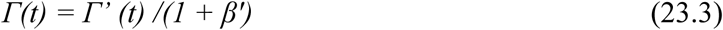

From the above we can see that for, *t*→*0*, we can write, *dx(t)/dt ∼ a(0) x(t)*, which would give the initial evolution to be, *x(t) ∼x(0) exp(a(0)t)= x(0)exp(t/τ*_*0*_) where *τ*_*0*_ *= 1/a(0*) is the typical time for initial exponential growth, in absence of quarantine or of lockdown.

The evolution equation (22) is a time dependent Riccati equation [6], whose quadrature form is known in the case when *a(t)* and *b(t)* are time independent. We shall solve the time independent case first, in Appendix I, from which we shall derive a formal expression for *x(t)* that can be used for different types of time-dependences of *a(t)* and *b(t*), as given in Appendix II.

It can be shown that Eq.(22) has a solution of the form, [*Appendix I* and *II* and refer to Eq (AII.15) therein.]

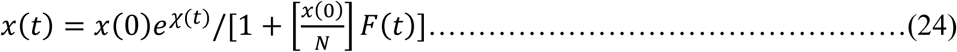

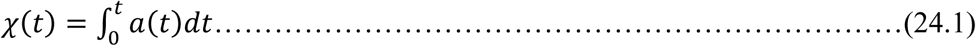

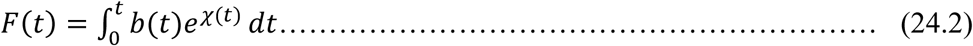

Expression (24) is an exact solution of the evolution equation for *x(t)*, in a formal sense. By considering the temporal evolution of *α(t), β(t)*, we can numerically evaluate *x(t)* by using (24). It can be seen qualitatively that

a. Infections grow and reach a saturation if for, *t* →*∞, χ(t)* →*∞*
b. Infection finally vanishes, if for, *t* →*∞, χ(t)* → *-∞*.

It can be seen from Eq.(22) that *x(t)* will surely decay as *t* →*∞, if for t* →*∞, a(t)* →*0, i*.*e. Γ’(∞) < γ(∞)*.

Further, quantification of the above formula can be obtained as follows. We find from (22) that in absence of recovery, the initial time scale of growth is [1/*Γ′(0)]*, that yields an initial “bare” (without recovery) time of *t*_*double*_*(0) = [ln(2)/Γ’(0)]*. Thus, to get a decay in *x(t)*, we must enforce, *Γ’(∞)=Γ′(0)Q(∞)=[ln(2)/ / t_double_ (0)] Q(∞) <γ(∞)*.

We shall present the graded lockdown model in the next section, to be followed by results.

### Model

We consider,

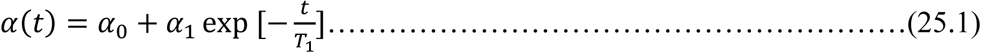

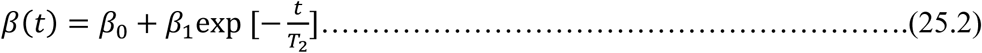

where, *0 ≤α(t), β(t)≤ 1*. The quantities *α(t), β(t)* are controlled externally. On the other hand, *τ*_*1*_ and *τ*_*2*_ are considered as constants which are decided by the nature of the disease. The parameters, *α*_*1*_, *β*_*1*_ can be positive or negative, depending upon whether *α(t), β(t)* are to rise or fall with time. In the case of rise, i.e. graded lockdown, *α*_*1*_, *β*_*1*_ have to be –ve. We can see,

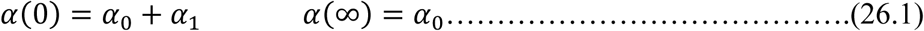

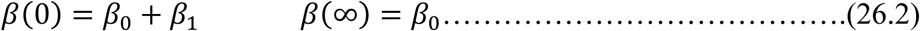

with *T*_*1*_ and *T*_*2*_ being the respective time scales for these changes. Since, initially, there is no quarantine, we must have, *α(0) = β*_*0*_*(0) = 0*, i.e. *α*_*1*_ *= -α*_*0*_, *β*_*1*_ *= -β*_*0*_, while, α_0_ and β_0_ are the respective final values of *α(t)* and *β(t)*.

Thus we have,

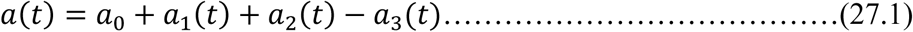

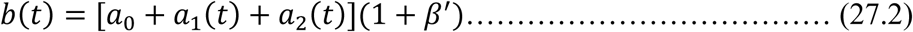

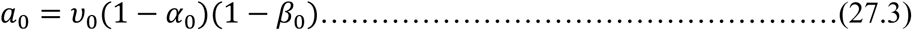

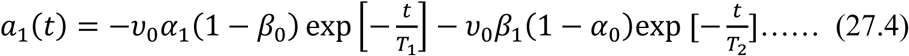

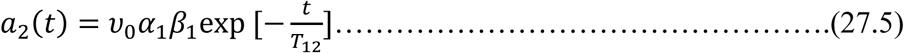

With

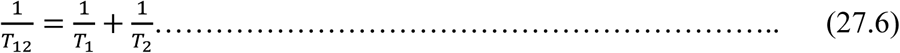

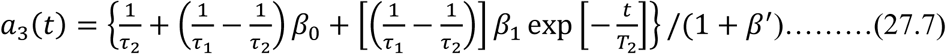

These finally yield,

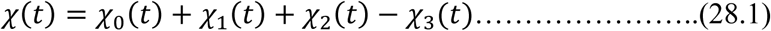

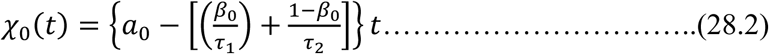

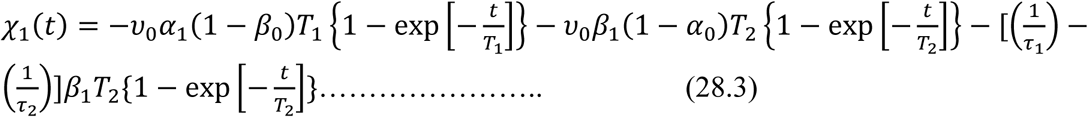

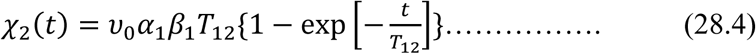

With realistic choice of these parameters, computation can be made.

## Results

In the present section we give our findings for six different wards of the Bruhat Bengalutu Mahanagara Palike (BBMP) which administers the city of Bengaluru. The city as a whole has a population 6.8×10^6^ living in an area of 771 sqkm, i.e. average population density is 8819 per sqkm. The population density in different wards varies widely being 113590 in Paadarayanapura (ward no. 135) and 1026 in Begur (ward no. 192) which are the two extremes. For computational purposes, the “sample” wards were chosen to represent different population densities. The values of the population, area, population density are given in Table 1. We have used *p*_*0*_ *= 2* meters to be the “infective length” which is also the necessary physical distancing length scale; the typical shoulder width of the population is chosen to be, *w= 0*.*4* meter. In spite of several inquiries we failed to get any estimate from physicians about the coughing and sneezing rates υ_0_ of infected persons. We chose, υ_0_ = *480* times per day, i.e. one (sneeze or cough) event per every *3* minutes, (this is the typical case for tuberculosis patients) but reduced it by a factor of *(1/2)* since people do not meet at night. We shall further consider, *φ(p*_*min*_,*p*_*max*_*) =1*.

**Table 1.**

| Sl No | Ward No. | Ward name | $N$ | $A$ | $\rho$ | $F$ | $\tau_{double}$ | $F$ | $\tau_{double}$ |
| --- | --- | --- | --- | --- | --- | --- | --- | --- | --- |
|  |  |  |  | Km<sup>2</sup> | Km<sup>-2</sup> |  | days |  | days |
| 1. | 65 | Kadumalleswara | 34053 | 1.36 | 25039 | 0.0741 | 2 | 0.0741 | 8 |
| 2. | 135 | Paadarayanaura | 35213 | 0.31 | 113590 | 0.0163 | 2 | 0.0004 | 8 |
| 3. | 189 | Hongasandra | 23058 | 2.16 | 10675 | 0.2500 | 2 | 0.0625 | 8 |
| 4. | 37 | Yashvanthpur | 35972 | 0.78 | 46118 | 0.0426 | 2 | 0.0100 | 8 |
| 5. | 92 | Shivajinagar | 35740 | 0.43 | 83116 | 0.0223 | 2 | 0.0057 | 8 |
| 6. | 85 | Doddanekundi | 22016 | 12.12 | 1817 | 1.0000 | 2 | 0.2555 | 8 |

We have put the recovery times for the infected patients to be: *τ*_*1*_ *= τ*_*2*_ *= 14* days-which are some typical values. The reduction factor *F* is not known though it is suggested by experts that if the infected person wears double layered cloth masks, there can be a reduction, with typical *F* ∼ *0*.*10* This can be reduced to *F ∼ 0*.*1×0*.*1 = 0*.*10*, if the infected person and the uninfected ones, both wear masks. In other words, we assume that everyone wears a mask, uniformly of same *F* value, in a given zone. In Table I, we display the values of *F* that are to be satisfied to make the initial doubling time to be 2 days and 8 days, in different BBMP wards.

### Bengaluru cases

For the quarantine-fractions *Q(t)*, we let α(t) rise from *α(0)= 0* to *α(∞) = α*_*0*_ and let *β(t)* rise from *β(0)= 0* to *β(∞) = β*_*0*_, with the same time scales of rise set at, *T*_*1*_, *T*_*2*_ *= 2* and *10*. The results are shown in Figs. 2-6, with the parameters being displayed in each figure caption.

**Figure 2:**
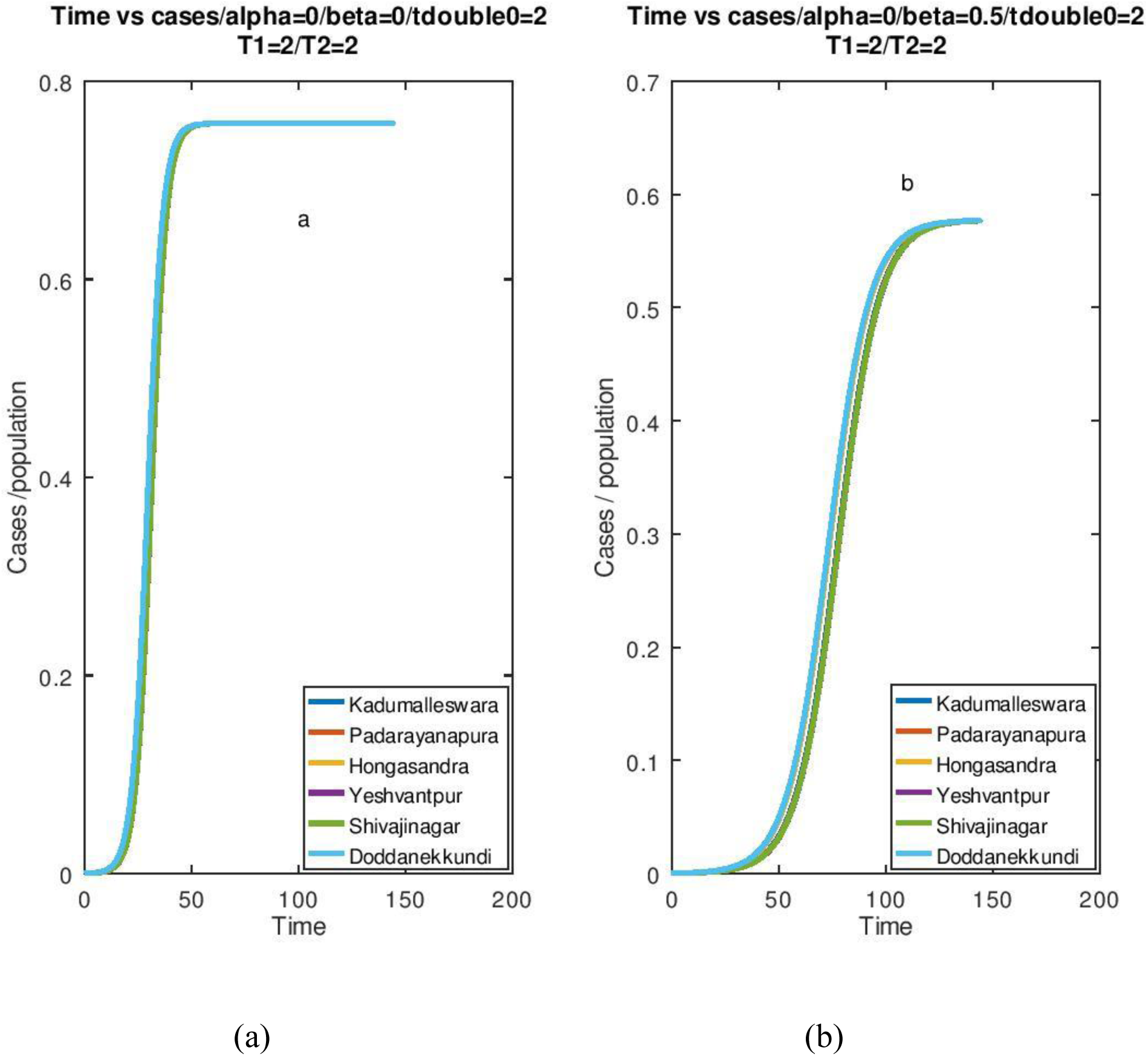
In both graphs *T*_*1*_*=T*_*2*_*=*doubletime0= *t*_*double*_ = *2 days*. In the graph (a) *α(t)=0* and *β(t) =0*. In graph (b) *α(t)=0* and *β(∞)=0*.*5*. The different curves in both plots are overlapping.

### Chikkaballapur cases

In the last part of this section we consider the case of 6 municipal towns, namely Bagepalli, Chikkaballapur, Gouribidanur, Chintamani, Gudibanda, Siddalaghata, with populations per sqKm being 5286,3422,5413,5210, 14751, 14330. In all the cases we have kept *F= 0*.*3513*, which makes the initial doubling time to be 2 days for Bagepalli. Accordingly, the initial doubling times for the towns are 2, 3.09, 1.95, 2.03, 0.72, 0.74 days respectively in the order in which the names of these places appear in the first sentence of the paragraph. Different parameters for these towns is given in Table 2 and the growth of *[x(t)/N]* for the different towns to follow, as given in Figs 7-10.

**Table 2.**
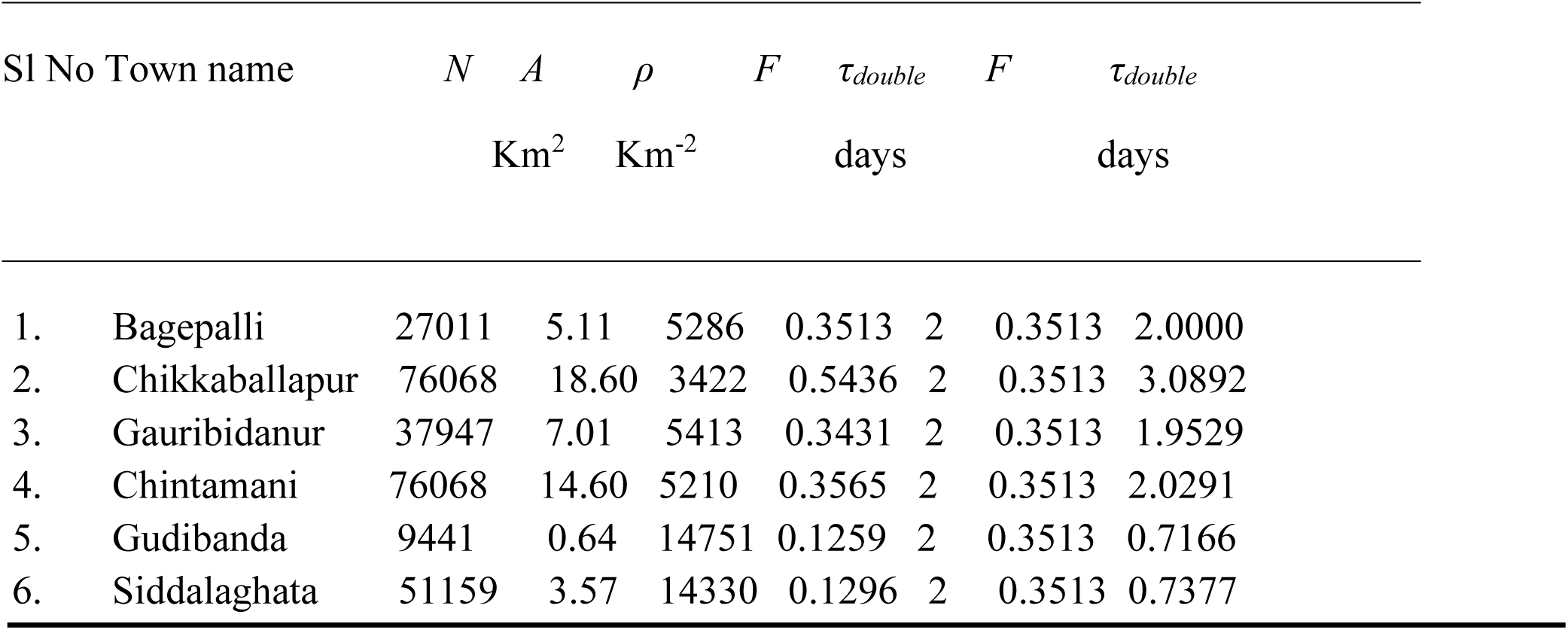

## Discussion

Figures 2-4 show the manner in which the cases vary. Here, we have considered *T*_*1*_*=T*_*2*_*=2* days, that is, a maximum time of growth of 2 days for the quarantining process to reach the saturation. The initial doubling time is also taken to be 2 days for all the wards by choosing *F* accordingly. Figure 2 shows a continuous increase in the number of cases which ultimately saturate. The 14 day time gap required for the natural process of cure is the only factor to decrease the number of cases from this saturation number. From figure 3, we see the decay in the number of cases when the lockdown reaches 79.6%-the critical point for the case. Thereafter, from figure 4, a decrease in the number of cases is seen for a stricter lockdown. Quarantine of healthy people appears to have only a marginal effect, if 90-99% of the infected people are already quarantined.

**Figure 3:**
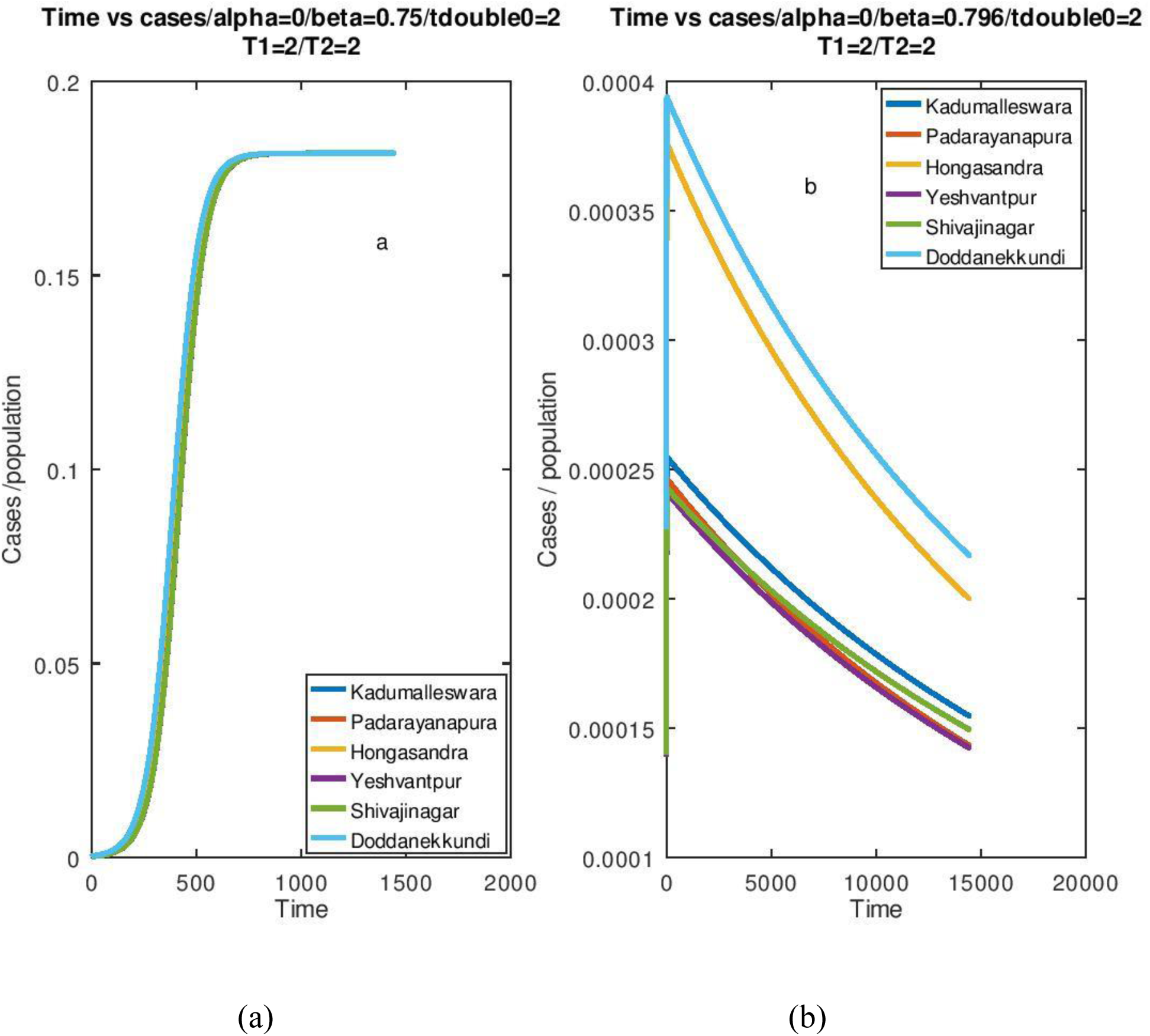
In both graphs *T*_*1*_*=T*_*2*_*=* doubletime0 = *t*_*double*_ =*2 days*. In the graph (a) *α(t)=0* and *β(∞)=0*.*75*. In graph (b) *α(t)=0* and *β(∞)=0*.*796*. The different curves in both plots show similar trend. In plot (a), the cases reach saturation but in plot (b), we find a decrease in the number of cases, although over a very long period of time.

**Figure 4:**
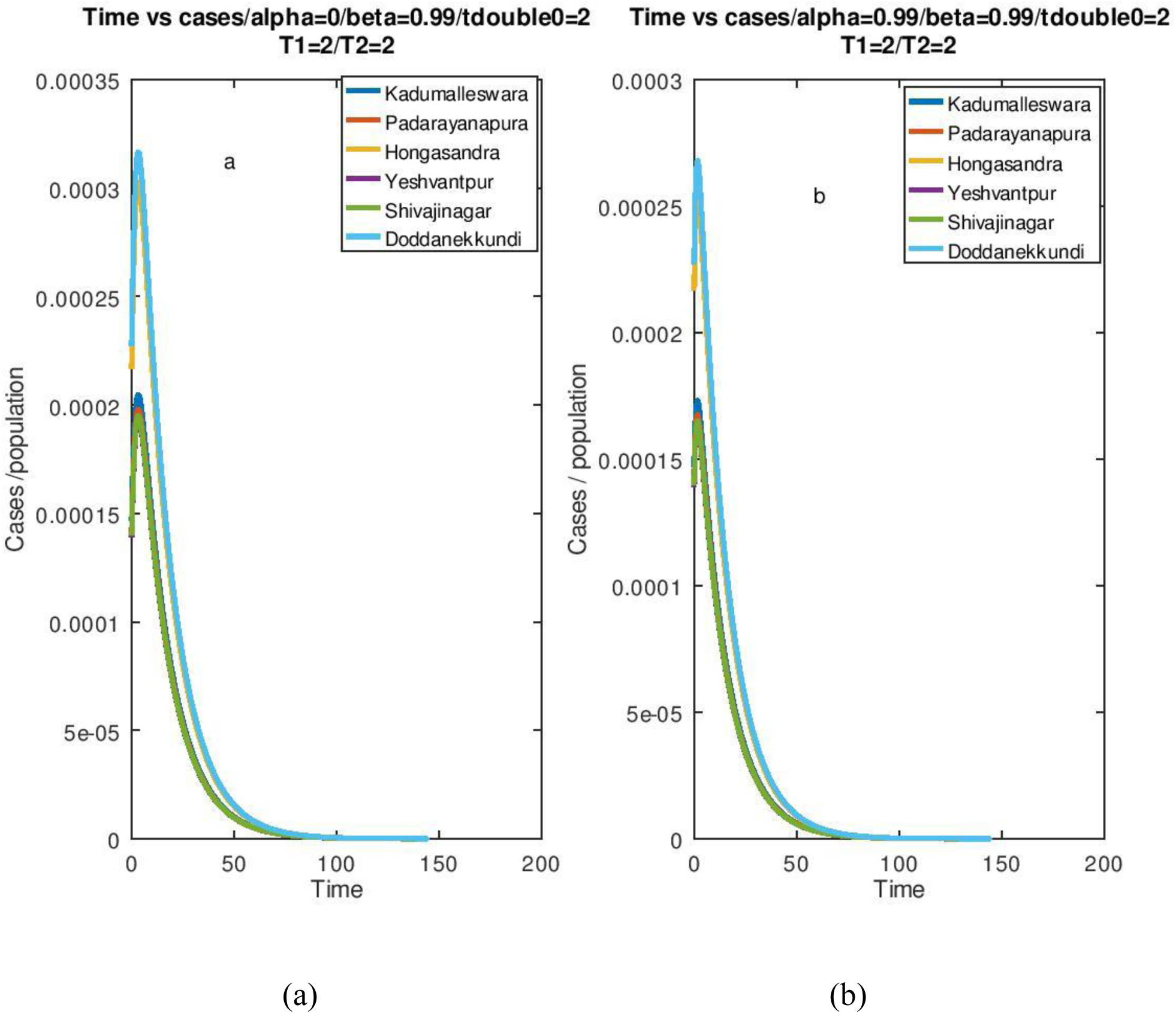
In both graphs *T*_*1*_*=T*_*2*_*=* doubletime0= *t*_*double*_ = *2days*. In the graph (a) *α(t)=0* and *β(t)=0*.*99*. In graph (b) *α(t)=0*.*99* and *β(t)=0*.*99*. The different curves in both plots show similar trend. Both plots show the cases going to zero in about 100 – 150 days.

For the above parameters, the decrease in the number of cases starts at =0.796, but, we find that it needs more than 20000 days to go to zero cases. This is to be expected since close to the “critical” or “transition” point, for saturation to decay, the time scale of decay would be very large. Obviously, this cannot be a practical solution. We thus need stricter quarantine protocol for containment. Figure 4 gives the situation for 99% lockdown quarantine. Here, it takes about 100 for the cases to drop to zero, after an initial rise. This means that all protection mechanisms and stipulated quarantining should be followed for at least for 100 days, to avoid the infection reappearing.

Figure 5 shows the situation when the initial doubling time is 8 days. Time available for lockdown has been raised to 10 days. In spite of these relaxed timescales, we find that the cases start to decrease at a low quarantine rate of just 50%. The decrease in fact starts at beta value of 0.17, or 17% of the infected people are kept under quarantine. This has a very important message. If the initial doubling time can be made slow, the virus’s spread can be controlled, even under instances of lockdown violation.

**Figure 5:**
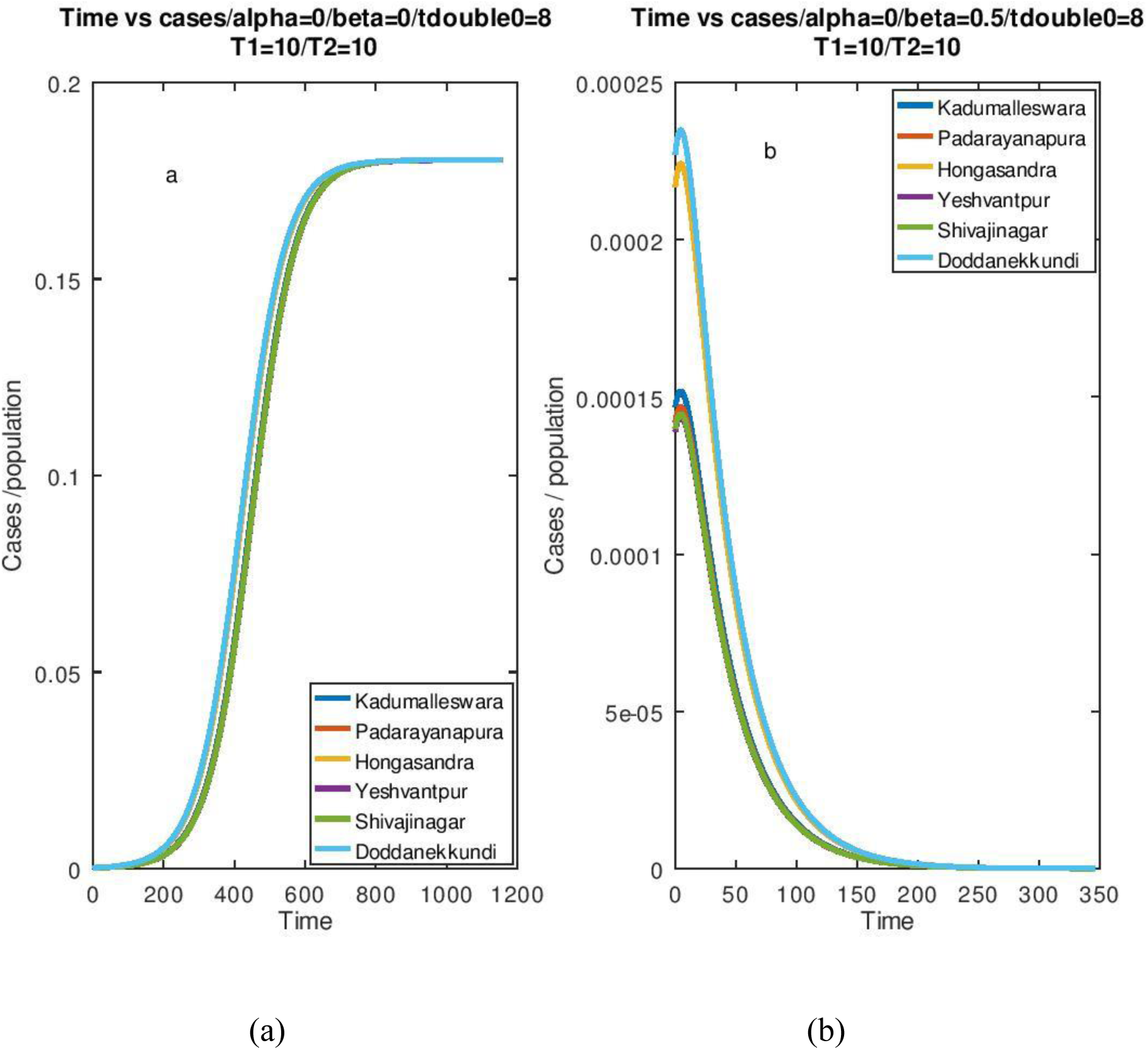
In both graphs *T*_*1*_*=T*_*2*_*=10 days* and doubletime0=*t*_*double*_ = 8 days. In the graph (a) *α(t)=0* and *β(t)=0*. In graph (b) *α=0* and *β=0*.*5*. The different curves in both plots show similar trend. In plot (a), cases reach saturation, whereas in plot (b), decrease in cases over time is seen.

Figure 6 shows the evolution of the doubling time for *α=0, β=0* and 0.5 and *T*_*1*_*=T*_*2*_=initial doubling time=2. As we know from figure 1, these represent conditions where the number of infected cases reach a saturation. As expected, the doubling time keeps increasing, as the infection spreads. Lockdown does increase the doubling time, but the aim is not increasing the doubling time, instead we need the cases to start decreasing. This can be achieved only if the lockdown of infected people is effectively enforced above a critical percentage.

**Figure 6:**
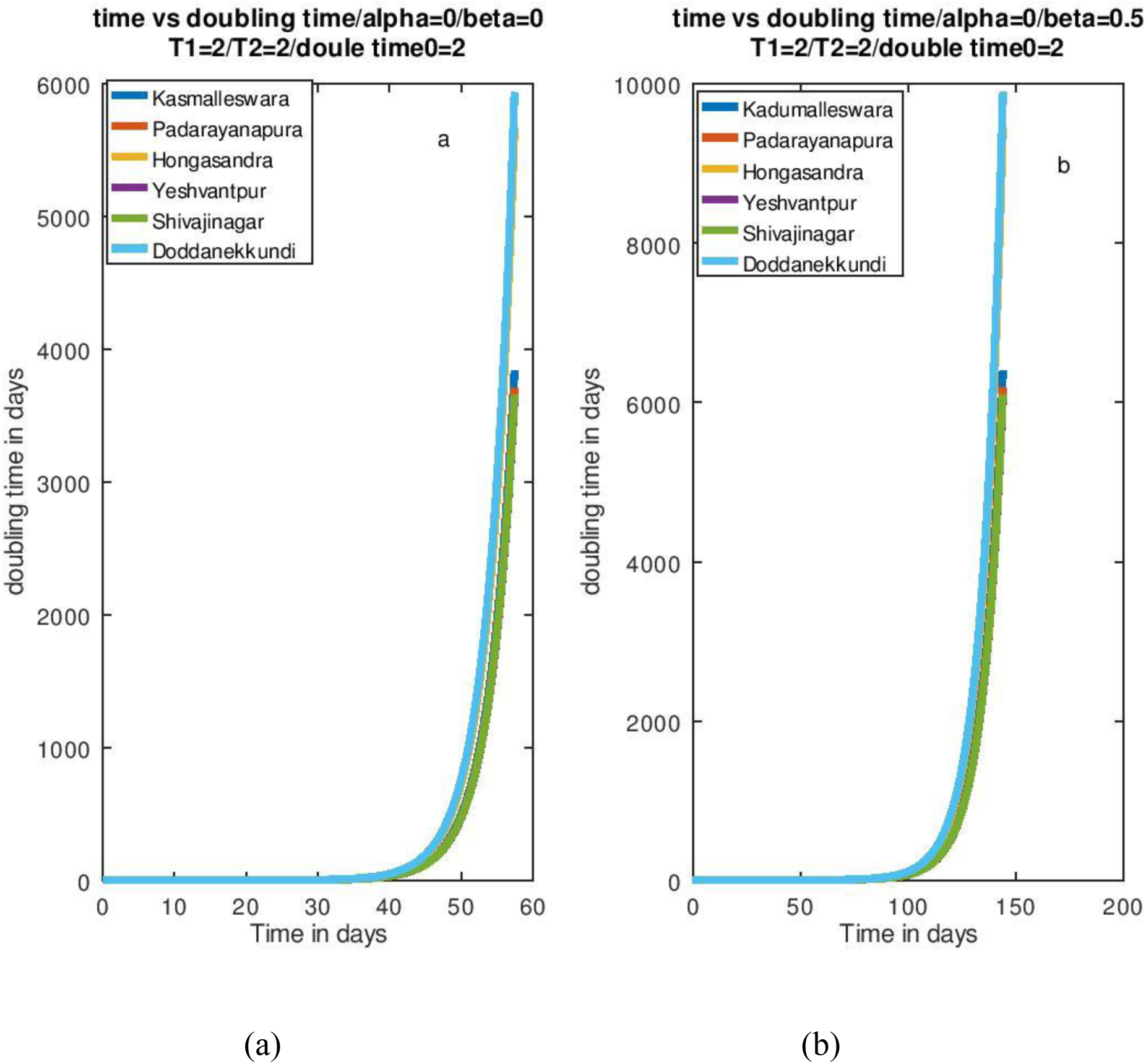
This graph shows the evolution of doubling time. In both graphs *T*_*1*_*=T*_*2*_*=*doubletime0= *t*_*double >*_*2*. In the graph (a) *α(t)=0* and *β(t) =0*. In graph (b) *α(t)=0* and *β(t)=0*.*5*. The different curves in both plots are overlapping.

Figures 7-10 show the growths for cases given in Table 2. Here, since the population densities are lower, the critical values can be reached for lower values of *β(∞)*. However, as in the case of BBMP wards, total lockdown does not have any significant effect, compared to what would be achieved in 99% of infected cases could be quarantined.

**Figure 7.**
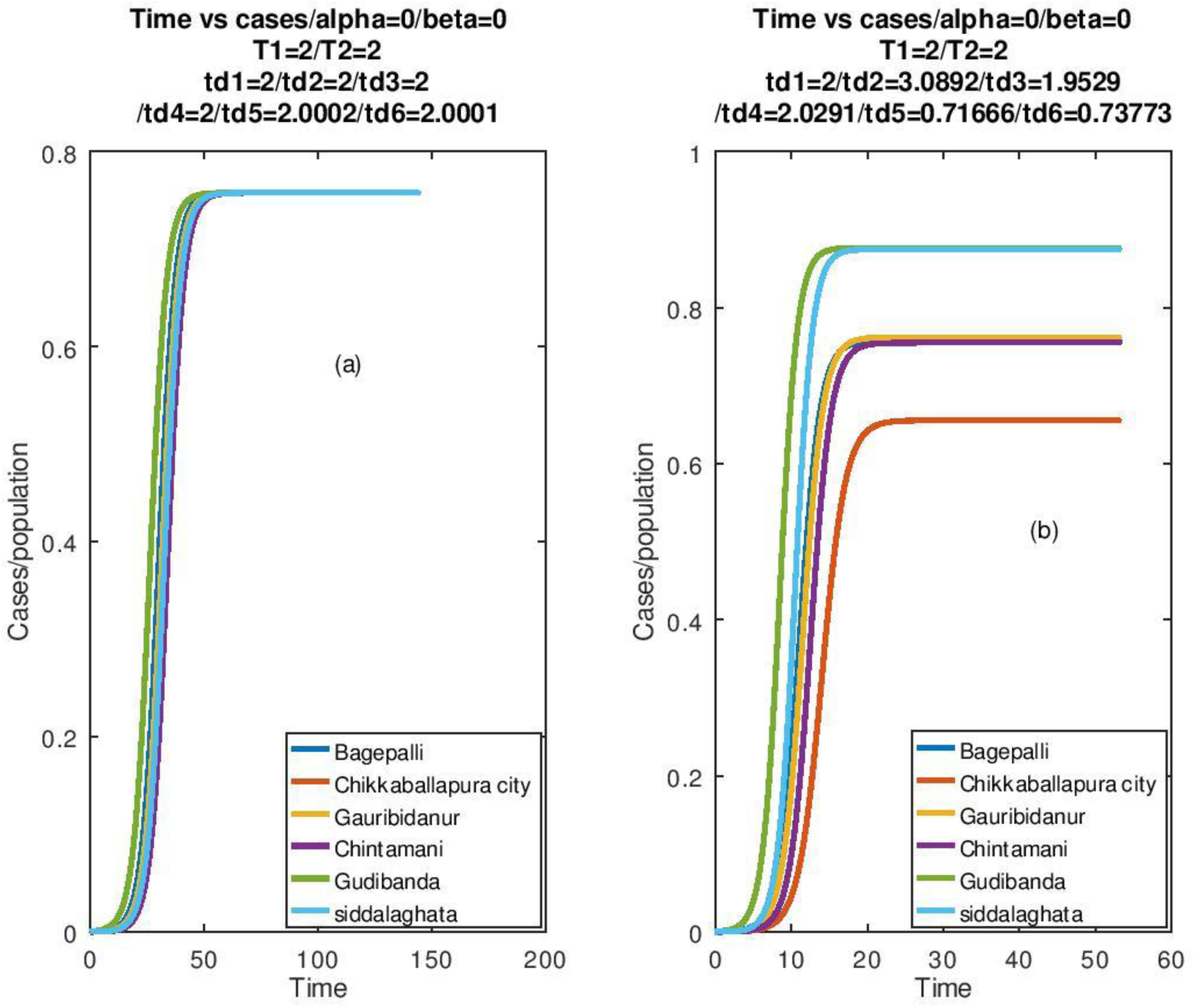
Growth curves for cases in Table 2. In subplot (a) the *F’*s were adjusted to give the initial doubling time for all cases to be 2 days keeping *α(t) =0= β(t)*. It is seen that all of them reach the same *[x(t)/N]* as *t*→*∞*. In subplot (b) *F* is kept fixed for all cases and we find that they saturate at different *[x(t)/N]*. The *t*_*d*_’s designate the doubling times for the different towns.

**Figure 8.**
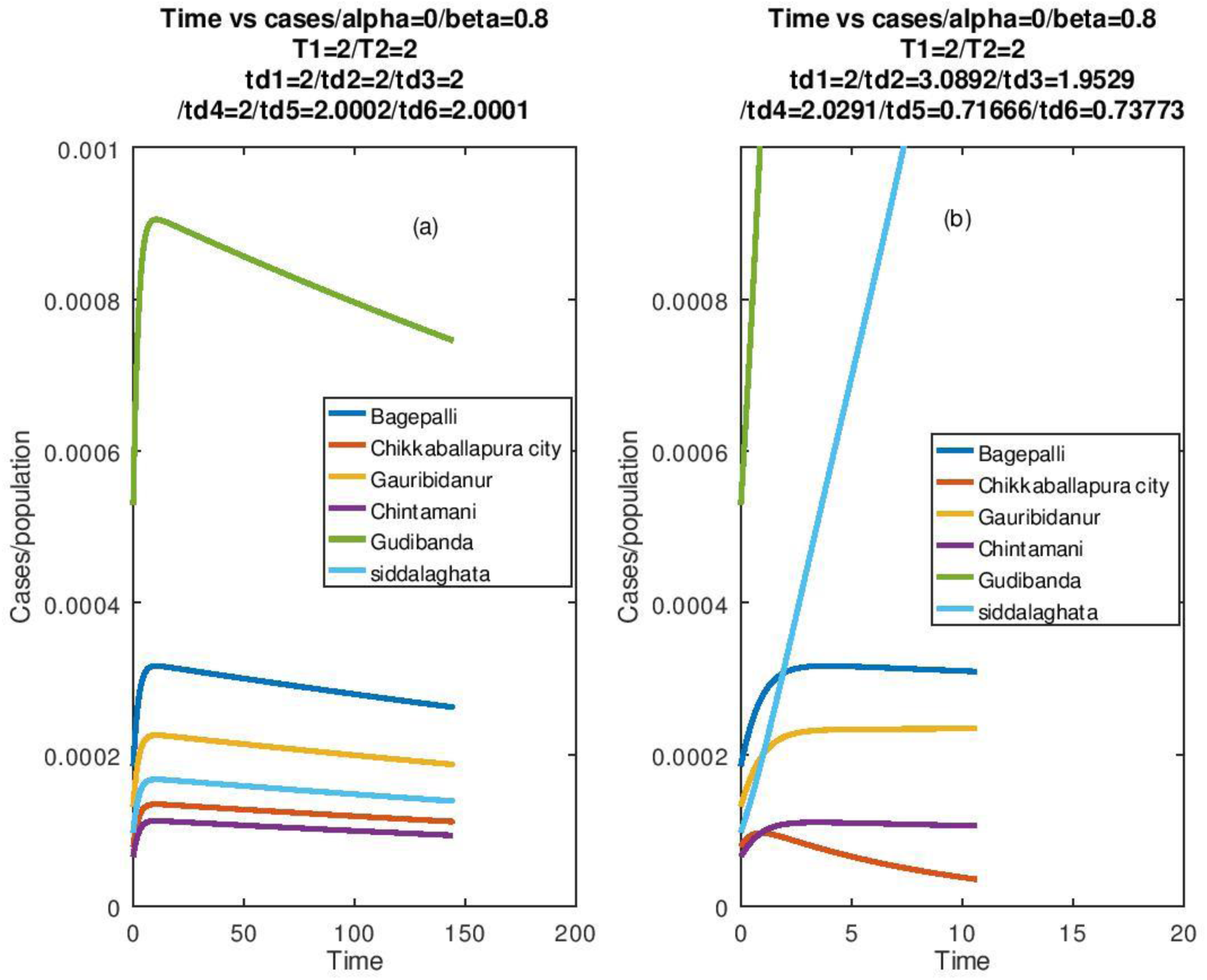
Growth curves for cases in Table 2. In subplot (a), the *F*’s were adjusted to give the initial doubling time for all cases to be 2 days keeping *α(t) =0* and *β(t)* rises from *β(0) =0* to *β(∞) =0*.*50*. It is seen that all of them decay. In subplot (b), *F* is kept fixed for all cases and we find that their growth curves differ, not only in detail but also in their qualitative nature. The *t*_*d*_’s designate the doubling times for the different towns.

**Figure 9.**
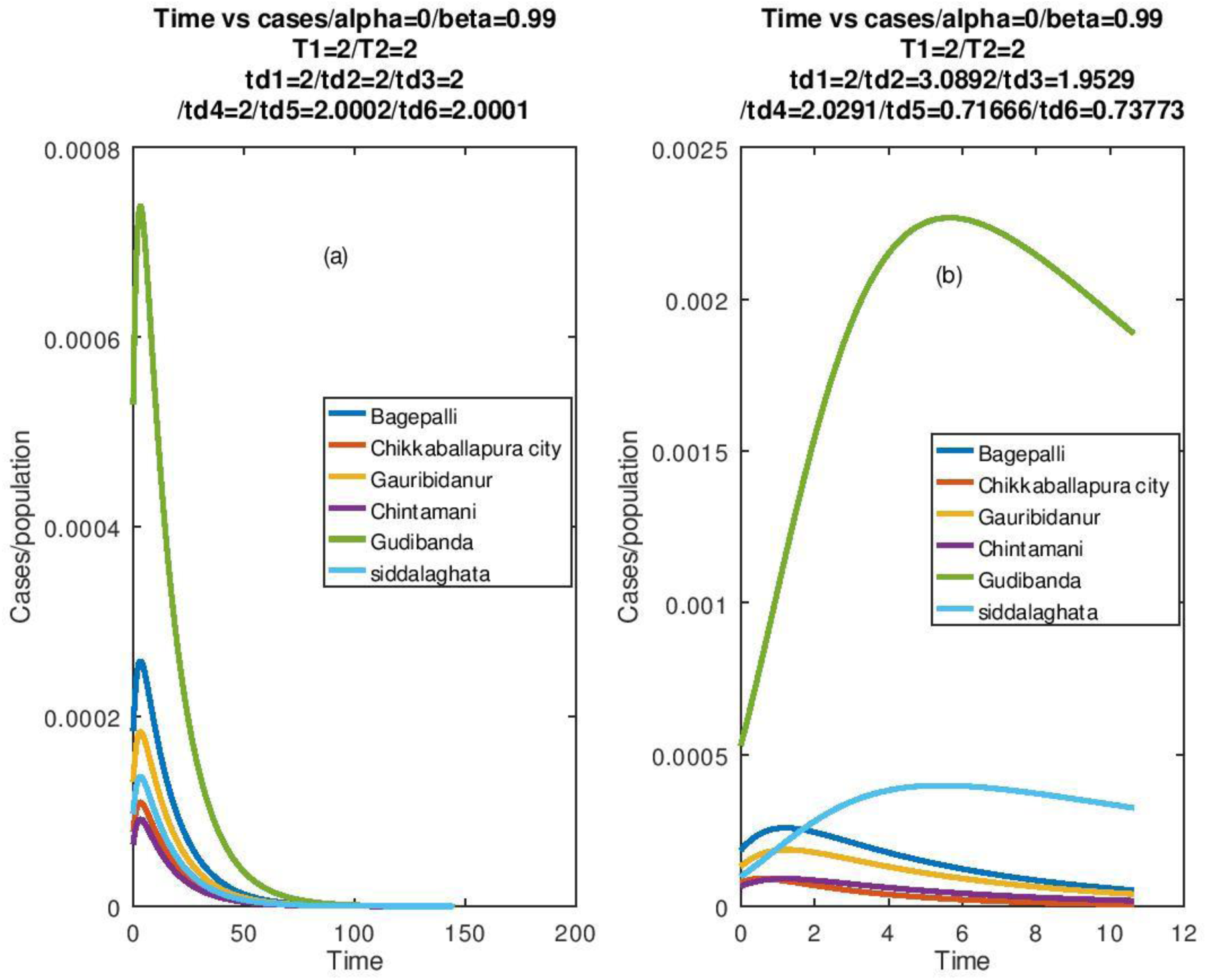
Growth curves for cases in Table 2. In subplot (a), the *F*’s were adjusted to give the initial doubling time for all cases to be 2 days keeping *α(t) =0* and *β(t)* rises from *β(0) =0* to *β(∞) =0*.*99*. It is seen that all of them decay. In subplot (b), *F* is kept fixed for all cases and we find that their growth curves differ, but qualitatively they decay after an initial growth. The t_d_’s designate the doubling times for the different towns.

**Figure 10.**
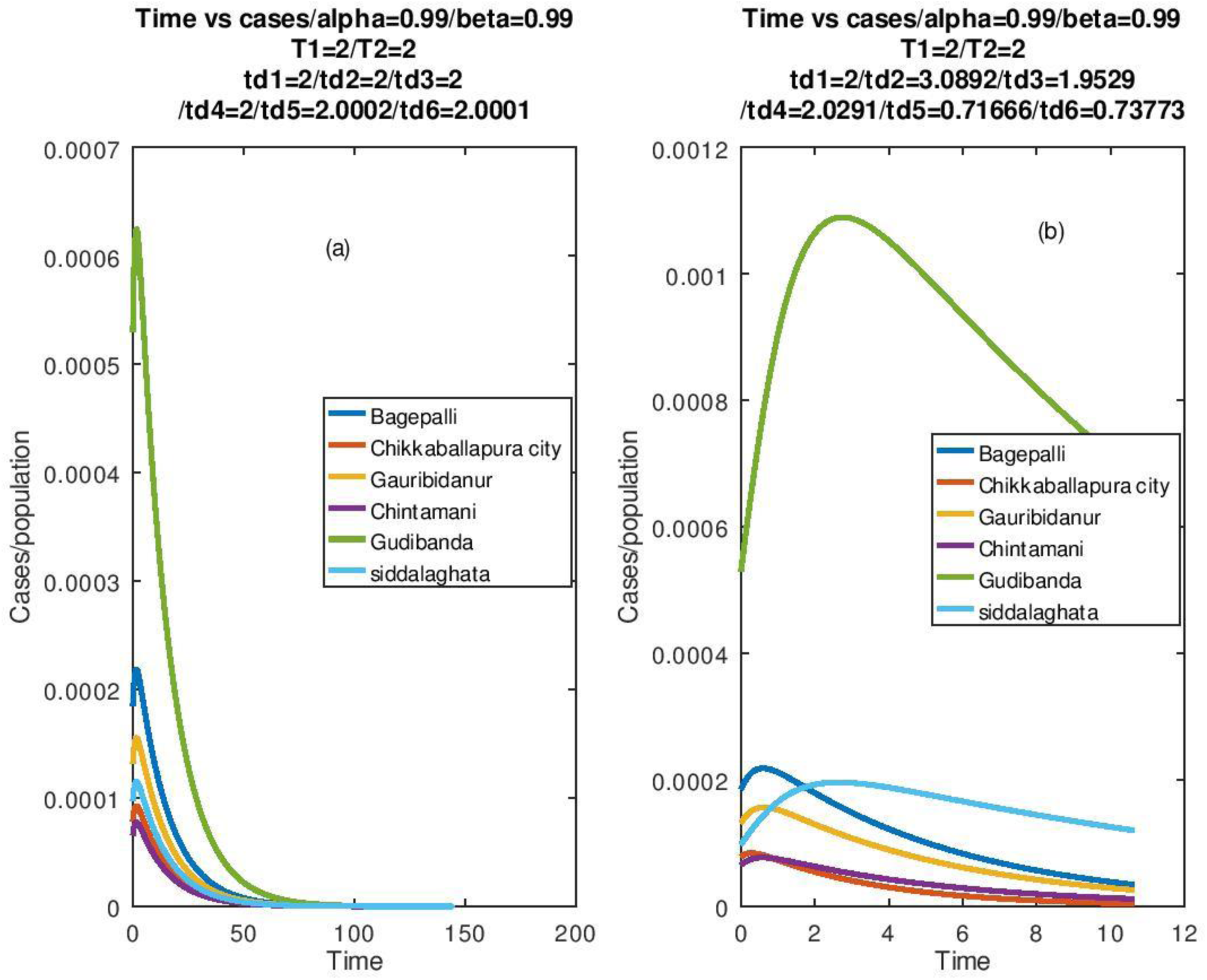
Growth curves for cases in Table 2. In subplot (a), the *F*’s were adjusted to give the initial doubling time for all cases to be 2 days keeping *α(0) = β(0) =0* and both *α(t), β(t)* rise to *α(∞) = β(∞) =0*.*99*. It is seen that all of them decay. In subplot (b), *F* is kept fixed for all cases and we find that their growth curves differ, but qualitatively they decay after an initial growth. It is to be noted that the difference between in the curves in Fig 7 and 8 have insignificant differences. The t_d_’s designate the doubling times for the different towns.

Most important step in any strategy would be compulsory use of masks, to make *F < 0*.*10*, as an essential step in social vaccination [7-8]. This has to be accompanied by quarantine of infected cases, as confirmed by medical practitioners. It is seen that if these steps are strictly followed, large-scale lockdowns fail to achieve any extra benefit. However, vigilance has to be a long-term one, covering several months [9] and on detection of an infected case, in any given area immediate surveillance and vigilant containment has to be executed [10]. Two of the authors are direct witness as to how containment without any lockdown can contain spread. This had happened in a residential apartment, only 100 meters from their residence. By immediately sealing off the building and alerting the local citizens of BBMP ward 65, spread was curbed very rapidly, so that no new case appeared in the next two weeks and also the infected patients recovered in two weeks by home quarantine. This had happened in a ward with relatively low density of population, the average being 25000 per square kilometer and the population density in this particular area of the ward is even smaller. For this purpose, more strict vigilance and quarantining schemes have to be kept ready for regions in the ward that have higher population density.

## Conclusion

### Above results lead us to the following conclusions concerning the ways to contain the spread of the epidemic

1. The total number will have an exponential rise initially but will come down subsequently if the proportion quarantined be high and quicker the process, higher is the gain, so that one has to try to make *T*_*2*_ comparable to the initial time scale of rise.
2. Enforce protection to make initial doubling time fairly long, 8 days (say).
3. For final infection number to be pushed to zero, the time scale of rise must be reduced urgently and brought down to a value which is comparable with the recovery time scale. If quarantining cannot achieve this on time, the number of infections may fall very slowly, infecting large number of people, before it falls to zero. Or else, it will saturate.
4. Ensure testing, confirming, quarantining ideally 90% to 99% of infected people. This process to be completed within 10 days. At places with lower density of population, a lower quarantine factor can work, but is not recommended since the PCR tests may also fail, in about 30% cases.
5. The decay time for the infections to fall to zero is determined by the recovery time of the patients. In this case, we have put τ_1_ = τ_2_ = 14 days. Hence, we see the time for fall to zero is nearly 100 to 150 days. Protection should be continued for at least 100 days to avoid infection reappearing. If ALL the above steps are satisfied, then quarantining uninfected people is of little consequence. It can give rise to social economic burden, which this theory cannot account for.
6. All the BBMP wards and towns of Chikkaballapur used in this numerical study show similar trends.
7. Need BBMP data and The Ministry of Health data with timeline of infection-spread.
8. Can apply to other epidemics and simultaneous outbreak of several epidemics.
9. False negative, which are a common feature, to be minimized, so that infected cases are not left un-quarantined.
10. For all the above cases, we have used the exponential and Gaussian cases and left out the Lorentzian (or Cauchy) cases, since estimates of *p*_*max*_ and *p*_*min*_ are not known.

It is hoped that these results would be useful for local planning.

## Data Availability

The data used in this study was taken from regular press releases by public health authorities and govt sources.

## Acknowledgement

The authors would like to thank Dr. Ali Reza Shakibafard of the Shiraz University of Medical Sciences, Shiraz, Iran. for making available the Covid-19 occurrence data in Shiraz. Two of us (Chatterjee and Vani) have benefitted from discussions with Dr. Prakash C Rao and Dr. A. Anil Kumar, private medical practitioners, from Yashvanthpur and Bagepalli respectively. Special thanks are due to Shri Manjubath Raju, Ward Councilor of Kadumalleswara, Ward 65 of Bengaluru and Shri Saurav Kumar, journalist from Muzaffarpur, Bihar. We also acknowledge encouragement from Prof T. Jacob John, formerly of the Christian Medical College, Vellore, Tamil Nadu, India.

## Appendix I

Solution for “a” and “b” being time independent

We first consider the case

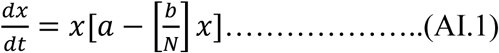

Or

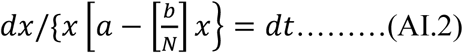

where a,b are constants.

Since

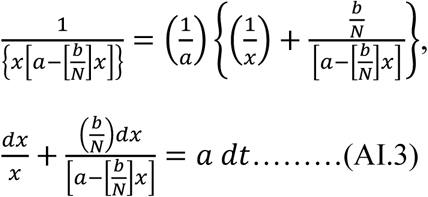

Then integrating (AI.3),

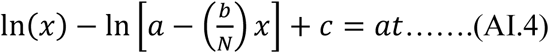

At t=0, we have x =x(0) so that,

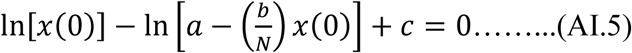

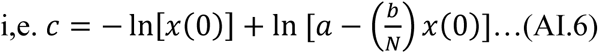

so that (AI.5) gives,

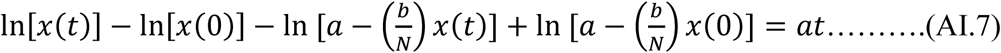

Thus,

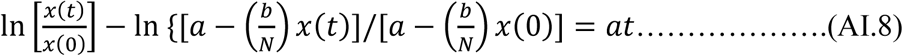

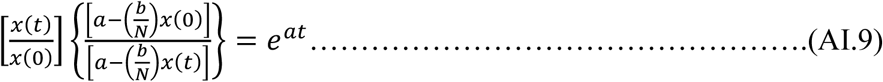

So that

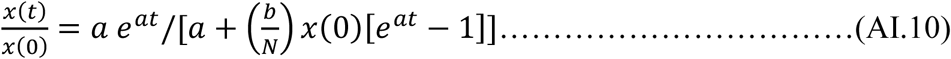

Thus, we see that at t=0, we have x(t) = x(0) and at t →*∞*, x(*∞*) = N(a/b) = N[1-(γ/Γ(1+β’))]/(1 +β’).

Thus, x(*∞*) becomes smaller as we make (γ/Γ) close to 1, i.e. make the time constants of recovery and growth to be close to each other. Suppose we make a =0, then x(t)/x(0) = 1 for all t. Thus, the growth can be stopped if we make a=0, which can be achieved if we make any of the factors [1-β(t)],, [1-α(t)] of Γ’ to be equal to zero. The condition [1-β(*∞*)] = 0, implies that ultimately there is a complete quarantine of all infected people, while [1-α(*∞*)] = 0 is satisfied when all non-infected people are quarantined. Since the growth can be stopped by making [1-β(*∞*)] = 0, i.e. quarantining infected people, it is obvious that nothing extra is gained by simultaneously making [1-α(*∞*)] = 0, i.e. complete lockdown.

## Appendix II Extension to time dependence case

This is done by using (33). Let us divide the time t into infinitesimally small intervals with t_n+1_ – t_n_ = ε →0. Then from (AI.9),

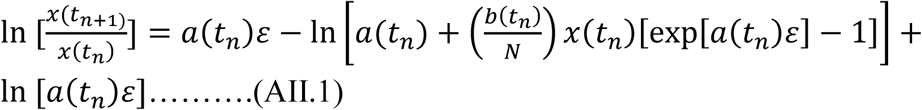

Since ε →0, we have [exp[a(t_n_)ε] −1] = a(t_n_)ε

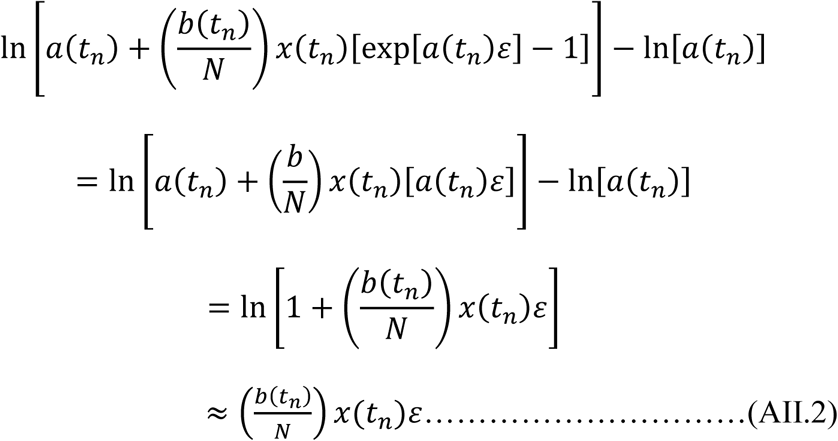

Thus substituting (AII.2) in (AII.1), we have,

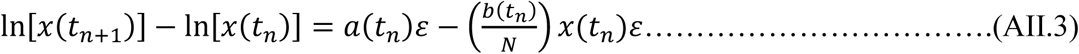

Thus, we can write successively,

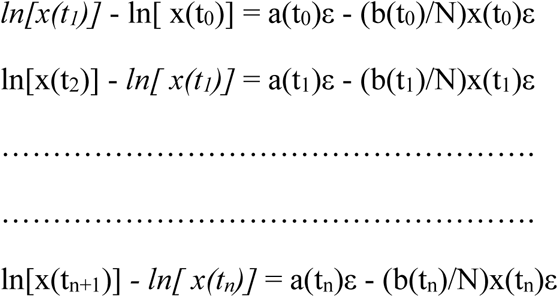

So that on adding and cancelling the terms in italics,

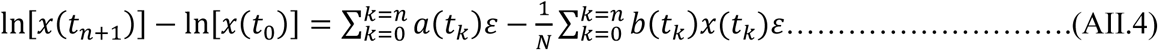

Since t_0_ = 0, we get on replacing the sum by an integral,

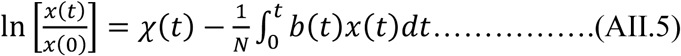

With

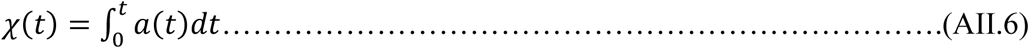

Equation (AII.5) is an integral representation of evolution of x(t), it is not a solution. We try to convert the integral equation to a differential equation by assuming the solution to be

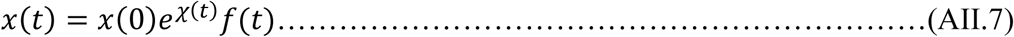

Then we have, on taking logarithm of both sides of (AII.6)

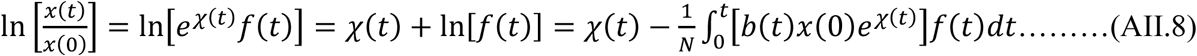

So that,

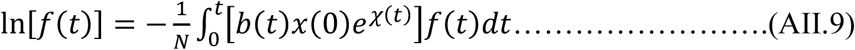

which is also an integral equation but on differentiating, w.r.t t, it can be converted to a differential equation, which reads,

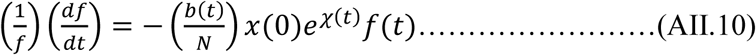

Thus,

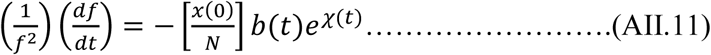

On integration, (AII.11) gives,

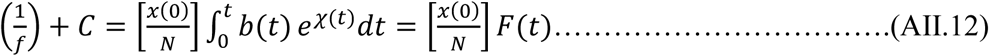

*where* 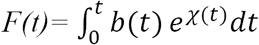

As an initial condition, we have that at *t= 0, f(0) = 1*, while the integral on the r.h.s. of (AII.12) vanishes, i.e. *F(0) = 0*. This gives us

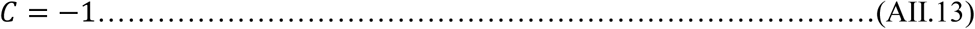

Hence,

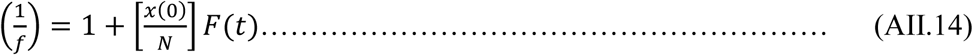

so that,

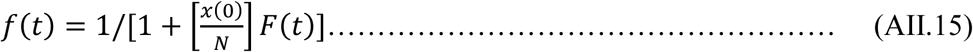

Thus, substituting (AII.15) in (AII.7), we have,

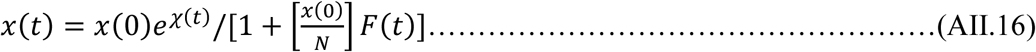

